# Identifying risk genes for embryo aneuploidy using ultra-low coverage whole-genome sequencing

**DOI:** 10.1101/2023.07.22.23292618

**Authors:** Siqi Sun, Mansour Aboelenain, Daniel Ariad, Mary E. Haywood, Charles R. Wageman, Marlena Duke, Aishee Bag, Manuel Viotti, Mandy Katz-Jaffe, Rajiv C. McCoy, Karen Schindler, Jinchuan Xing

## Abstract

**Background:** Aneuploidy, the state of a cell containing extra or missing chromosomes, frequently arises during human meiosis and is the primary cause of early miscarriage and maternal age-related in vitro fertilization (IVF) failure. IVF patients exhibit significant variability in aneuploidy rates, although the exact genetic causes of the variability in aneuploid egg production remain unclear. Preimplantation genetic testing for aneuploidy (PGT-A) using ultra-low coverage whole-genome sequencing (ulc-WGS) is a standard test for identifying and selecting IVF-derived embryos with a normal chromosome complement. The wealth of embryo aneuploidy data and ulc-WGS data from PGT-A has potential for discovering variants in paternal genomes that are associated with aneuploidy risk in their embryos.

**Methods:** Using ulc-WGS data from ∼10,000 PGT-A biopsies, we imputed genotype likelihoods of genetic variants in parental genomes. We then used the imputed variants and aneuploidy calls from the embryos to perform a genome-wide association study of aneuploidy incidence. Finally, we carried out functional evaluation of the identified candidate gene in a mouse oocyte system.

**Results:** We identified one locus on chromosome 3 that is significantly associated with maternal meiotic aneuploidy risk. One candidate gene, *CCDC66,* encompassed by this locus, is involved in chromosome segregation during meiosis. Using mouse oocytes, we showed that CCDC66 regulates meiotic progression and chromosome segregation fidelity, especially in older mice.

**Conclusions:** Our work extended the research utility of PGT-A ulc-WGS data by allowing robust association testing and improved the understanding of the genetic contribution to maternal meiotic aneuploidy risk. Importantly, we introduce a generalizable method that can be leveraged for similar association studies using ulc-WGS data.

## Background

Cells containing an abnormal number of chromosomes, a condition called aneuploidy, is the most common genetic abnormality in human embryos and the leading genetic cause of miscarriage and *in vitro* fertilization (IVF) failure [1]. Maternal age is well documented as a risk factor for producing aneuploid gametes. However, the propensity to produce aneuploid embryos varies substantially even among mothers of a similar age [1–5]. Recently, variants in several genes related to control of chromosome segregation have been implicated in contributing to aneuploidy risk [5–8]. However, many identified variants only contribute to the aneuploidy risk in a small number of patients and most of these studies have limited sample sizes. Additional efforts are needed to fully understand the genetic contribution to the aneuploidy risk in populations.

Currently, the most effective treatment of infertility is IVF, where eggs are surgically retrieved after controlled ovarian stimulation and fertilized in a petri dish, with subsequent embryo selection and transfer back to the uterus [9, 10]. Preimplantation genetic testing for aneuploidy (PGT-A) was developed as an approach to improve IVF outcomes by prioritizing euploid embryos for transfer, based on the inferred genetic constitution of an embryo biopsy [11, 12]. PGT-A with ultra-low-coverage whole-genome sequencing (ulc-WGS) performed on trophectoderm cells isolated from blastocyst-stage embryos has provided a rich resource of aneuploidy measurements. However, due to the low coverage of the genome (< 0.01x genome coverage per embryo biopsy), the genotype information encoded therein is rarely used for genetic studies in understanding infertility [12, 13].

Genome-wide association studies (GWAS) have revolutionized the field of complex disease genetics over the past decade by identifying genotype-phenotype associations based on testing millions of genetic variants across the genomes [14, 15]. For genetic variants showing strong disease association, further fine-mapping and gene prioritization approaches proceed to identify variants that causally impact the traits [16, 17]. This integrated approach has identified risk loci for many diseases and traits, such as susceptibility to viral infections and type 2 diabetes [18, 19]. Applying GWAS approach to PGT-A data would help identify additional genetic risk factors to embryo aneuploidy.

Here we describe an integrative approach to identify candidate variants through retrospective analysis of ulc-WGS-based PGT-A data. After combining data from sibling embryos and imputing variant dosages, we conducted a GWAS to identify candidate genes. Our analysis identified one genomic region that is associated with embryo aneuploidy risk on chromosome 3. Functional interpretation of the variants suggested that the candidate variants are causal eQTLs for *CCDC66.* Validation experiments in mouse oocytes showed that CCDC66 depletion was associated with higher aneuploidy rates.

## Methods

### Dataset description

PGT-A data were obtained from IVF cases between 2017 and 2019 at CCRM Fertility. One IVF cycle with at least three embryos tested was included for each patient. IVF cycles with maternal age >=43 years were excluded from the analysis because eggs used in these cycles were from egg donors of unknown age. Embryos underwent trophectoderm biopsy on day 5, 6, or 7 postfertilization, followed by PGT-A using the Illumina VeriSeq PGS kit and protocol, which entails sequencing on the Illumina MiSeq platform (36-bp single-end reads) (Illumina, USA). Chromosome copy numbers from each embryo biopsy were inferred using the Illumina BlueFuse Multi Software suite in accordance with the VeriSeq protocol, as described before [20]. Each embryo was then noted as “euploid” or “aneuploid” based on the chromosome copy number.

The aneuploidy rate for each IVF cycle was determined with the formula described previously [7, 8, 21]:

*aneuploidy rate = (no. of aneuploid embryos)/(total no. of embryos tested)*.

### Sequencing alignment and variant calling

PGT-A sequencing files with < 150,000 reads were considered low quality and excluded. After filtering, sequencing files from each IVF cycle were combined into a single file for analysis. The sequencing reads were aligned to the human reference genome (GRCh38) with bwa-mem (v 0.7.17) [22] and converted to the BAM format using samtools (v 1.13) [23]. Ancestry inference was performed using LASER (V2.0) as previously described [20, 24, 25]. Briefly, principal component (PC) space was defined based on the 1000 Genomes project reference samples. Sequencing samples were then projected onto the space using a Procrustes approach implemented in LASER. Samples were assigned to superpopulations (African [AFR], Admixed American [AMR], East Asian [EAS], European [EUR], and South Asian [SAS]) based on genetic similarity to the 1000 Genomes reference panel.

Genotype likelihoods (GLs) were computed with bcftools (v 1.13) [26] for each sample at all variable positions of the reference panel (1000 Genomes 30x on GRCh38, https://www.internationalgenome.org/home). Imputation and phasing in the form of GLs were performed using GLIMPSE [27]. Specifically, GLIMPSE refines the GLs by iteratively running genotype imputation and haplotype phasing with a Gibbs sampling procedure to produce consensus-based haplotype calls and genotype posteriors at every variant position [27]. With imputed data, each variant site was filtered based on the following criteria: imputation score >= 0.2, minor allele frequency (MAF) >= 5%. After filtering, the imputed genotype dosages of each patient were calculated and used in the association test:

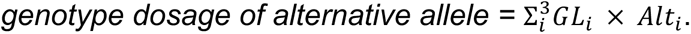

Given the known reference and alternative alleles, *GL_i_* of three possible genotypes (*i.e.*, homozygous reference, homozygous alternative, and heterozygous) were multiplied by the number of alternative alleles of each genotype (*Alt_i_*).

For MAF correlation analysis, the population MAF for each variant was extracted from two reference panels, the 1000 Genomes Projects (http://ftp.1000genomes.ebi.ac.uk/vol1/ftp/data_collections/1000G_2504_high_coverage/working/20201028_3202_phased/) and the Genome Aggregation Database (gnomAD) (v3.1) [28]. Correlations of MAFs between our imputed data and reference panels were calculated with Pearson correlation coefficient (R).

### Association test and eQTL analysis

For the association test, a quasibinomial generalized linear regression model (GLM) was iteratively fit for each variant using the function glm() in R as follows: *glm(data, formula = cbind (aneuploid_embryos_numbers, euploid_embryos_numbers) ∼ age + ancestry_PCs + single_SNP_dosage, family = “quasibinomial”)*

At each iteration, a single nucleotide polymorphism (SNP) dosage was tested. Maternal age and top four ancestry PCs inferred using LASER were included as covariates. The resulting p values were visualized using a Manhattan plot and checked using a QQ plot with the R package GWASTools (version 1.44.0) [29]. The significant variants were determined using a significance threshold p value ≤ 2×10^−8^ and Benjamini-Hochberg false discovery rate (FDR) ≤ 0.05. Haplotype structure surrounding significant loci was visualized with Locuszoom (http://locuszoom.org/) [30].

The Genotype-Tissue Expression (GTEx) project includes genotypes, gene expression, and histological and clinical data from 54 non-diseased tissue sites across nearly 1,000 individuals [31]. The eQTL information from GTEx (https://www.gtexportal.org/home/eqtlDashboardPage, access date: 06/30/2022) was used to determine the candidate variants’ potential association with expression of nearby (*i.e.*, cis) gene.

### Mice and oocyte collection and maturation

C57BL/6 mice (6–10 weeks and 9 months of age) (Jackson Laboratory, USA) were used. Mice were housed with a constant temperature and a standard 12 h light/12 h dark cycle in the animal facility at Rutgers University (NJ, USA). All animal experiments performed in this study were approved by the Rutgers IACUC (protocol #201702497) and followed guidelines set by the National Institutes of Health. For oocyte collection, mice were primed with pregnant mare serum gonadotropin (PMSG, Lee Biosolutions, #493-10) two days before collection. Prophase I-arrested oocytes were collected as described before [32] in minimum essential medium (MEM) (Sigma, #M0268) with 2.5 µM milrinone (Sigma, #M4659) to prevent spontaneous meiotic resumption. The oocytes were then incubated in Chatot, Ziomek, and Bavister (CZB) media without milrinone, in 5% CO_2_ at 37^°^C for the desired time of maturation, depending on the meiotic stages to be evaluated (0h for Prophase I, 5h for Pro-metaphase I, 7 h for Metaphase I, and 16 h for Metaphase II).

### Knockdown of CCDC66 in mouse oocytes

To deplete CCDC66, we used the Trim-away strategy [7, 33, 34]. Rabbit anti-CCDC66 antibody (Bethyl Laboratories, #A303-339A) and control IgG antibody (Merck Millipore, #12-370) were purified using Amicon Ultra 0.5-ml Centrifugal Filter (Merk Millipore, #UFC5003096). pGEMHE-Cherry-TRIM21 (Addgene, #105522) or pGEMHE-mEGFP-mTrim21 (Addgene, #105519) were linearized with Asc I (New England Biolabs, #R0558S,) and *in vitro* transcribed using a T7 mMessage mMachine Kit (Ambion, #AM1340). Prophase I-arrested oocytes were co-microinjected with the fluorescently tagged *Trim21* cRNA and with either rabbit anti-CCDC66 antibody (0.5 mg/ml) or IgG antibody (0.5 mg/ml) in the control group. Injections were performed using a Xenoworks digital microinjector (Sutter Instruments) in MEM supplemented with 2.5 μM milrinone. The oocytes were incubated in milrinione-containing CZB media for at least 3 hours in 5% CO_2_ at 37^°^C before starting meiotic maturation by washing out the milrinone and culturing in CZB medium. Oocytes were fixed at Metaphase I stage (7 h post milrinone washout) and immunostained to evaluate CCDC66 knockdown efficiency.

### Antibodies and immunofluorescence

The following antibodies were used: rabbit anti-CCDC66 antibody (1:50, Bethyl Laboratories, A303-339A), mouse anti-α-tubulin ((B-5-1-2) Alexa Fluor 488) (1:100, Invitrogen, 322588), and human anti-centromeric antigen (ACA) (1:30, Antibodies Incorporated, 15-234). These secondary antibodies (1:200) were used: donkey-anti-rabbit Alexa Fluor 568 (Life Technologies, A10042) and goat-anti-human Alexa Fluor 633 (Life Technologies, A21091).

Immunofluorescence was performed as previously described [35]. Oocytes were fixed with 2% paraformaldehyde (PFA) (Sigma-Aldrich, P6148) in phosphate-buffered saline (PBS) at room temperature for 20 min. The fixative was then washed out by incubating the oocytes in blocking buffer (0.3% BSA containing 0.01% Tween-20 in PBS) three times for 10 min. Oocytes were then permeabilized in PBS containing 0.2% Triton-X-100 for 20 min and blocked in blocking buffer for 10 min. Primary antibody incubation was performed by incubating the oocytes overnight at 4^°^C (CCDC66) or 1h at room temperature (ACA) in dark, humidified chamber, followed by three washes of 10 min each in blocking solution. Then oocytes were incubated in secondary antibody for 1 h in a dark humidified chamber, followed by three washes of 10 min each in blocking buffer. Finally, oocytes were mounted in 10 μl of Vectashield containing 4, 6-Diamidino-2-Phenylindole, Dihydrochloride (DAPI) (Life Technologies, D1306).

### *In situ* chromosome counting

As described previously [36, 37], the microinjected prophase I-arrested oocytes from young and old mice were matured in CZB media without milrinone in a humified incubator (5% CO_2_, 37^°^C) for 16 h until they completed meiosis I and arrested at metaphase of meiosis II. Then, eggs were cultured for at least 2 h in 100 µM Monastrol (Sigma #M8515) to collapse the spindle and facilitate the separation of the chromosomes. The eggs were fixed with 2% PFA in PBS for 20 min and permeabilized in PBS containing 0.2% Triton X-100 for 20 min. Eggs were stained with ACA antibody to detect centromeres and DAPI to detect DNA. Normal chromosome counts for a mouse egg is 20 pairs of sister chromatids; any deviation of this number was considered an aneuploid egg. Chromosome counting was performed with Image J software (NIH) using cell counter plugins.

### Imaging

Images were acquired with Leica SP8 confocal microscopes equipped with a 40×, 1.30 NA oil immersion objective or a 63×, 1.40 NA oil immersion objective. For each image, optical z-sections were obtain using 0.5µm step with zoom of 4.5. For comparison of pixel intensities, the laser power was kept constant for each oocyte in an experiment. All oocytes in the same experiment were processed at the same time.

## Results

### Project overview, patient cohort, variant calling, and ancestry inference

To identify genomic loci associated with aneuploidy in the embryos of IVF patients, we analyzed embryo biopsy sequences collected from the PGT-A procedure (Fig. 1). The dataset included 10,011 embryo biopsies from 1,467 IVF cycles. After removing data from egg-donors and low-quality (< 150,000 reads), 9,357 embryo biopsies from 1,373 cycles remained, with maternal age ranging from 23 to 42 years (median = 35) (Fig. 2A). To improve the coverage for analysis, we pooled all sequenced embryos from each IVF cycle. Because embryos in a cycle are equivalent of full siblings, this combined file captured both maternal and paternal genomes. After pooling, the median coverage of each patient was 0.056× (Fig. S1A). As expected, the mean coverage per patient was linearly associated with the number of sequenced embryo biopsies (Fig. S1B).

**Fig. 1.**
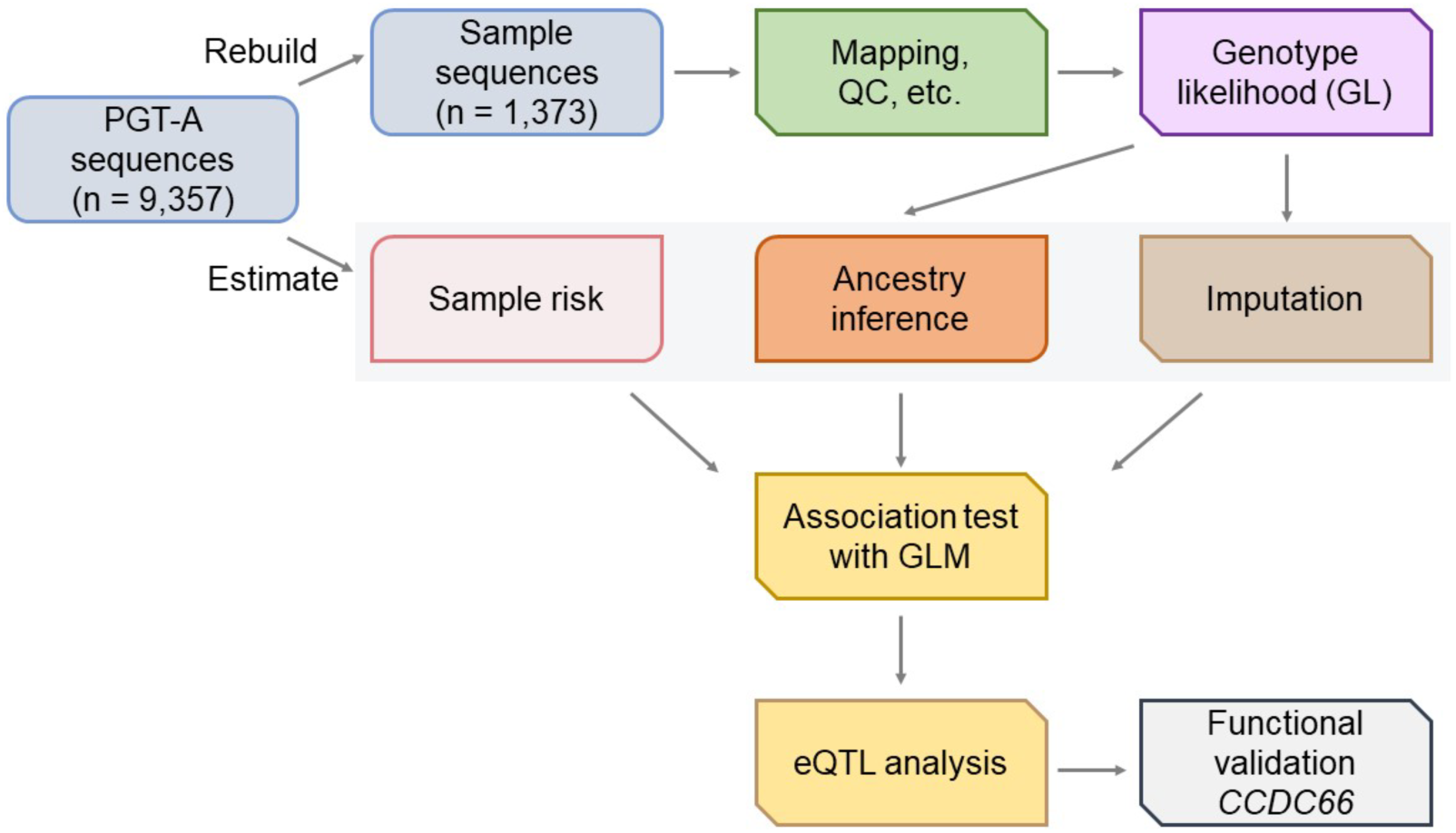
Overall strategy for PGT-A data analysis. For each IVF cycle, the number of aneuploidy and euploidy embryo biopsies were determined using PGT-A. ulc-WGS data from PGT-A for each IVF cycle were combined for analysis. Genotype likelihood and dosage were imputed for each variant and ancestry of the samples was inferred using the genotype likelihood. Association tests were performed between the imputed genetic variants and the aneuploidy rate. eQTL analysis was used to determine the candidate gene associated with the top variants from the association test. CCDC66 was selected for functional studies using a mouse oocyte model.

**Fig. 2.**
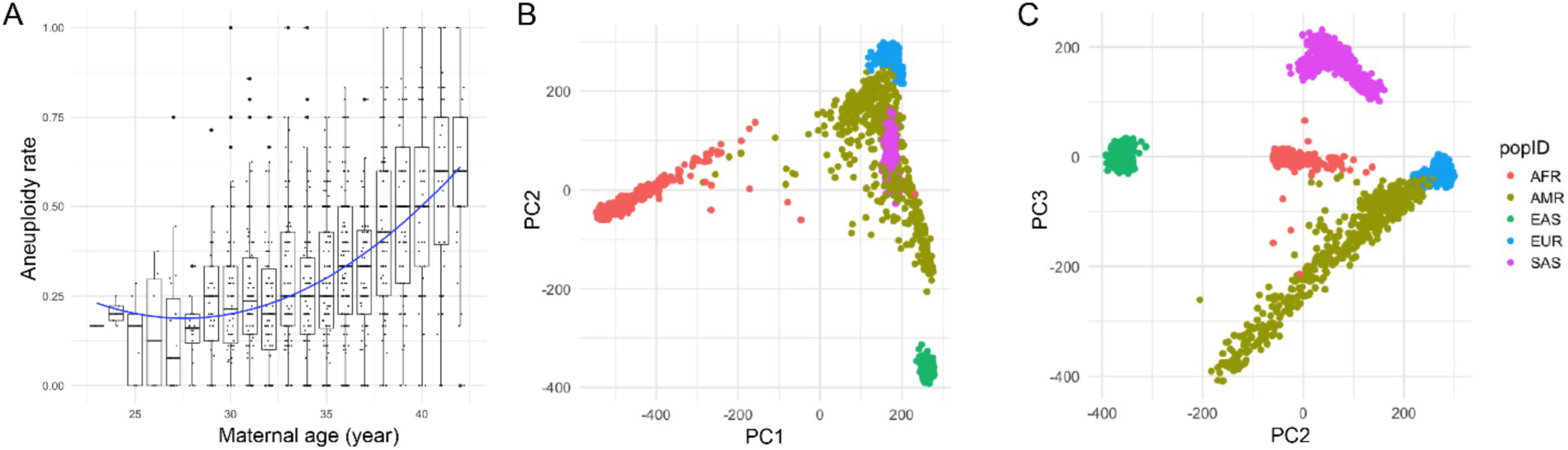
Phenotypic characterization and ancestry inference of the patients. **(A)** Aneuploidy rate versus age. x-axis: patient age in years, y-axis: blastocyst aneuploidy rate. **(B, C)** Ancestry inference using the PGT-A data. Principal component axes (PC1 and 2 in B, PC2 and 3 in C) were defined based on analysis of 1000 Genomes reference samples and colored according to superpopulation annotations (African [AFR], Admixed American [AMR], East Asian [EAS], European [EUR], South Asian [SAS]). PGT-A samples were then projected onto these axes using a Procrustes approach with LASER [24, 25].

We next performed ancestry inference based on the sequence data using the program LASER. Our analysis revealed a diverse patient cohort, consistent with the demographic composition of the local population (Fig. 2B, 2C). Specifically, according to the superpopulation reference panel defined by the 1000 Genomes Project [38], 788 samples (57.4%) have genetic similarity with European reference samples, 223 (16.2%) with Admixed American reference samples, 168 (12.2%) with African reference samples, 143 (10.4%) with South Asian reference samples, and 52 (3.8%) with East Asian reference samples.

Using the program GLIMPSE, we identified variants and performed GL imputation across the sample cohort (see Methods for details). A total of 10,740,080 variants were imputed, among which 4,353,993 variants had INFO scores >= 0.2 (Fig. S2A). After selecting variants with >= 5% MAF, 2,549,983 variants remained (Fig. S2B). After imputation, MAFs of imputed variants in our sample were highly correlated with large population databases: the 1000 Genomes (R = 0.95, p < 2.2e^−16^, Fig. S3A) and the gnomAD (R = 0.97, p < 2.2e^−16^, Fig. S3B).

### Genome-wide association analysis for aneuploidy

To identify aneuploidy risk loci, we next investigated the association between aneuploidy rate and genotype dosage for each variant using a GLM, incorporating four ancestry PCs and the maternal age as covariates (see Methods for details).

Three SNPs on chromosome 3 reached genome-wide significance for association with aneuploidy at the level of p <= 2×10^−8^ and FDR ≤ 0.05 (Fig. 3A; Table 1, Table S1). The QQ plot did not show strong inflation of the test statistics (Fig. 3B), suggesting that confounding factors, such as population structure, were generally controlled. The three significant SNPs were located in *ERC2* (ELKS/RAB6-Interacting/CAST Family Member 2), which has not been reported as associated with maternally-derived aneuploidy (Fig. 3C). Within the locus, the three significant SNPs are in strong linkage disequilibrium with each other (Table 1). The top SNP, rs12495172 (chr3-55959628-G-A), is located in intron 12-13 of *ERC2*. The mean depth of coverage of the 1 Mbp window covering the significant variants had a median of 0.066 among all samples, comparable to 0.055 for the entire chromosome (Fig. S3C, Table 1). As indicated by the positive beta values (*e.g.*, 0.079 for the rs12495172), the alternative allele of each significant variant in *ERC2* is positively associated with aneuploidy rate.

**Fig. 3.**
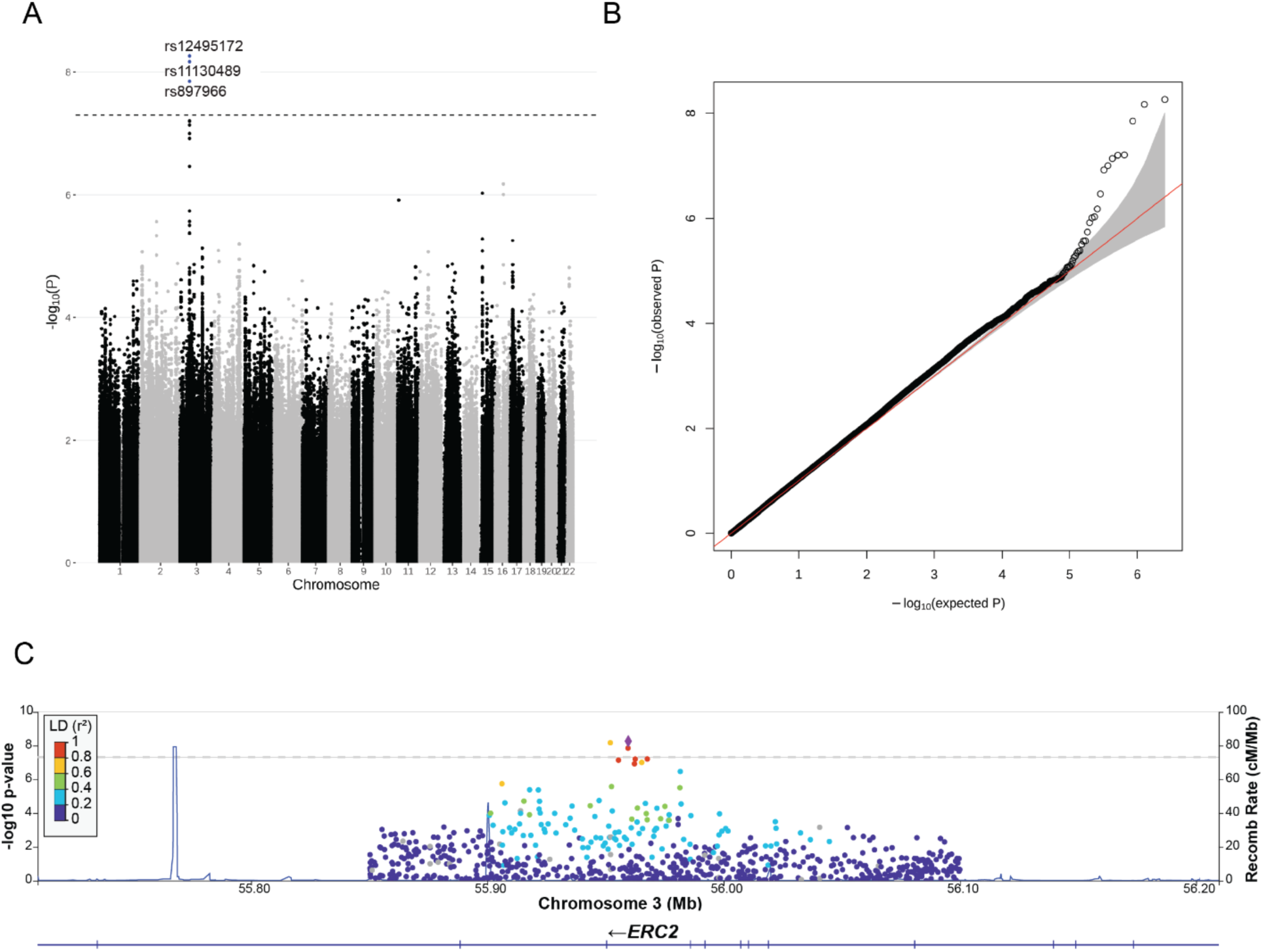
Association test for aneuploidy. **(A)** Manhattan plot depicts p values of association tests of each SNP versus the aneuploid embryo count. The rs numbers of the three significant SNPs on chromosome 3 are labeled. **(B)** QQ plot describes the distribution of observed p values versus those expected under the null hypothesis. **(C)** Locus Zoom plot denotes lead SNP (rs12495172) and SNPs around the region of *ERC2* gene. The lead SNP is shown in purple diamond and the heat map shows the linkage disequilibrium (LD) between the lead and nearby SNPs. Recombination rates are plotted as blue lines.

**Table 1.** SNPs associated with embryo aneuploidy.

| ID | Chr | Position | INFO | AF | AF<br>gnomAD | AF<br>1KG | p | FDR | beta | LD | Distance |
| --- | --- | --- | --- | --- | --- | --- | --- | --- | --- | --- | --- |
| 3:55959628:G:A | 3 | 55959628 | 0.237 | 33.8% | 39.4% | 40.8% | 5.48E-09 | 8.67E-03 | 0.079 | - | - |
| 3:55952031:C:T | 3 | 55952031 | 0.244 | 32.8% | 38.9% | 39.4% | 6.80E-09 | 8.67E-03 | 0.084 | 0.563 | 7597 |
| 3:55959515:A:G | 3 | 55959515 | 0.234 | 35.0% | 42.3% | 44.1% | 1.41E-08 | 1.20E-02 | 0.085 | 0.567 | 113 |
| 3:55967692:G:C | 3 | 55967692 | 0.242 | 34.8% | 41.6% | 43.0% | 6.23E-08 | 3.10E-02 | 0.082 | 0.559 | 8064 |
| 3:55962557:C:T | 3 | 55962557 | 0.245 | 34.2% | 41.8% | 43.1% | 6.30E-08 | 3.10E-02 | 0.078 | 0.613 | 2929 |
| 3:55955490:C:G | 3 | 55955490 | 0.239 | 35.0% | 42.8% | 44.4% | 7.29E-08 | 3.10E-02 | 0.079 | 0.621 | 4138 |
| 3:55965325:A:G | 3 | 55965325 | 0.247 | 32.8% | 38.1% | 38.8% | 9.96E-08 | 3.63E-02 | 0.080 | 0.559 | 5697 |
| 3:55962257:T:C | 3 | 55962257 | 0.235 | 34.7% | 39.8% | 41.0% | 1.20E-07 | 3.83E-02 | 0.083 | 0.618 | 2629 |

INFO: IMPUTE info quality score AF: alternative allele frequency AF gnomAD: alternative allele frequency in the gnomAD project AF 1KG: alternative allele frequency in the 1000 Genomes project beta: regression coefficient LD: linkage disequilibrium ( $r^2$ ) between the SNP and the top SNP (3:55959628:G:A) Distance: the distance (bps) between the SNP and the top SNP (3:55959628:G:A)

Next, we aimed to identify the candidate genes associated with the top variants. A previous study showed that variants discovered by GWAS are more likely to affect the expression of nearby genes, (*i.e.*, as expression quantitative trait loci, eQTLs) and the altered expression can ultimately influence the phenotypic trait [39]. Therefore, integrating GWAS with gene expression data can facilitate candidate gene prioritization [17]. To determine the effect of the top SNPs on nearby gene expression, we examined eQTL signals using GTEx project data. The GTEx data suggested that alternative alleles of the top variants were associated with reduced expression of a nearby gene *CCDC66* (Coiled-Coil Domain Containing 66) in two tissues (thyroid and tibial nerve, see Fig. 4A as one example). There was no eQTL signal for other genes, including *ERC2*. Therefore, we selected *CCDC66* as the candidate aneuploidy risk gene, whose reduction in expression may be associated with increased aneuploidy rate. As indicated by the positive beta values (*e.g.*, 0.079 for the rs12495172), the alternative alleles of the significant variants were positively correlated with aneuploidy rate (see Fig. 4B as one example).

**Fig. 4.**
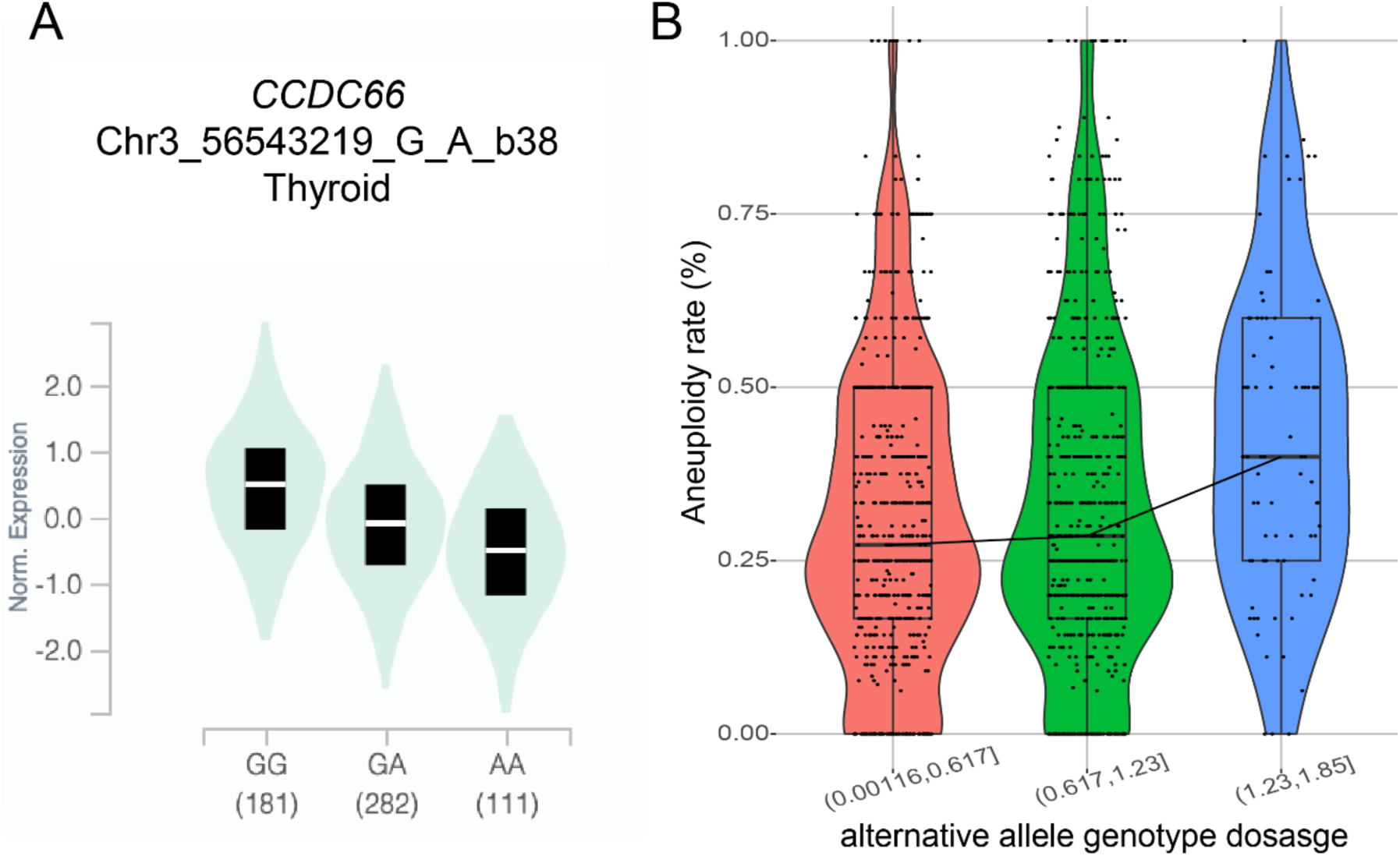
Gene prioritization for casual variants and genes. **(A)** GTEx eQTL of the lead significant variants rs12495172 (chr3-55959628-G-A) on *CCDC66* expression. The decreased expression of *CCDC66* correlates with alternative alleles of rs12495172. **(B)** The genotype dosages of the alternative allele of rs12495172. The alternative allele genotype dosages in samples were divided into 3 bins of roughly equal size. The aneuploidy rates among the samples were positively correlated with the alterative allele genotype dosages.

### CCDC66 regulates meiotic progression and chromosome segregation fidelity

*CCDC66* encodes a microtubule-associated protein that regulates microtubule nucleation and organization during cell division [40, 41]. In mitosis, CCDC66 regulates centrosome maturation via recruitment of core pericentriolar material (PCM) proteins and microtubule organization via its cross-linking activity [40].

To determine the role of CCDC66 in meiosis, we evaluated expression and localization of the protein during mouse oocyte meiotic maturation via immunostaining of oocytes fixed at different meiotic stages (Fig. 5A). We detected CCDC66 in Prophase I-staged oocytes with slight enrichment in the nucleus. In pro-Metaphase I and Metaphase I oocytes and in Metaphase II eggs, CCDC66 was enriched around the spindle (Fig. 5A). This localization pattern suggested a requirement of CCDC66 during mouse oocyte meiotic maturation.

**Fig. 5.**
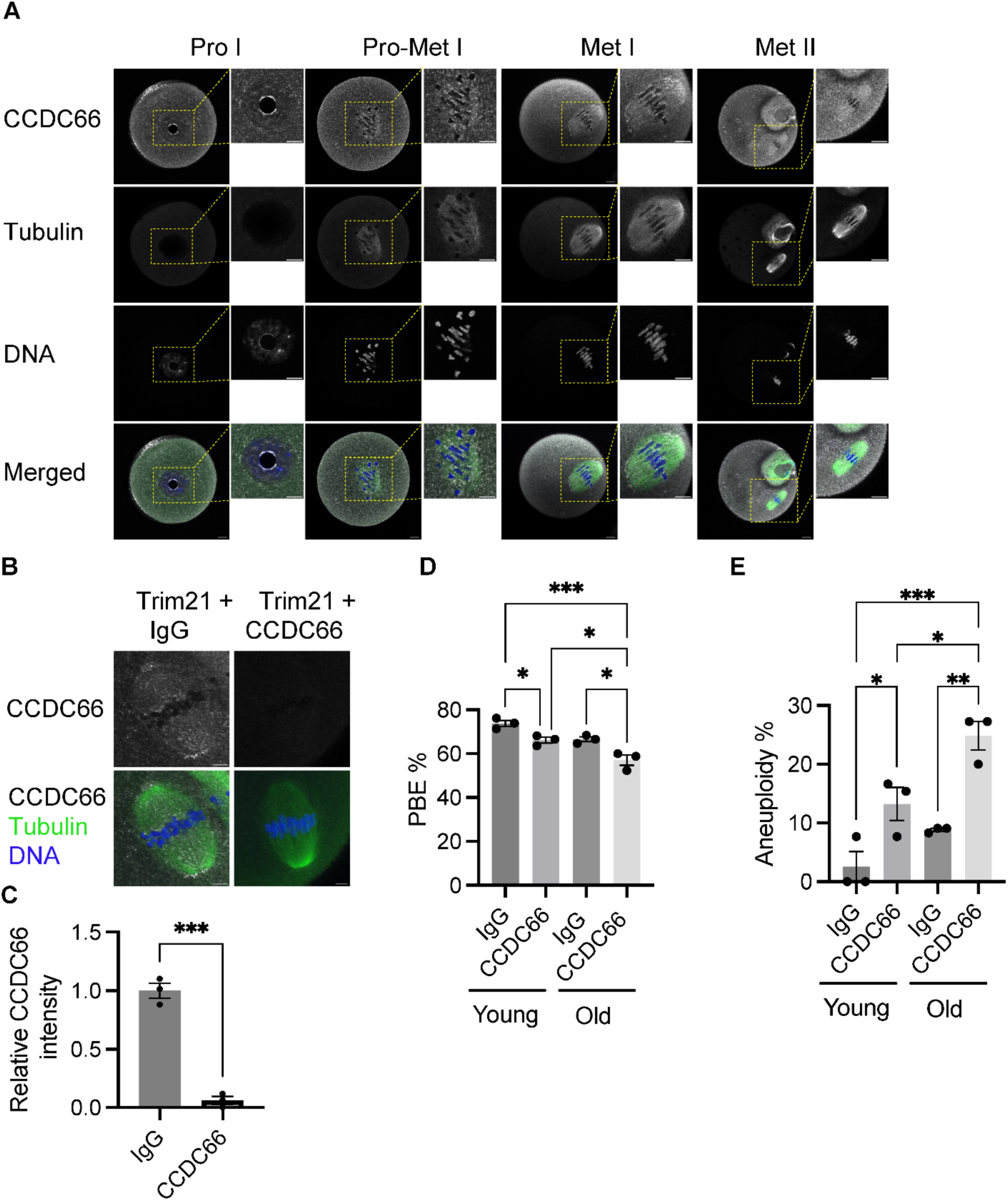
CCDC66 is required for meiotic progression and production of euploid eggs. **(A)** Expression and localization of CCDC66 in different stages of mouse oocyte meiotic maturation. Oocytes were stained with anti-CCDC66 antibody (gray) at (0, 5, 7, and 16) hours of maturation corresponding to Prophase I, Pro-metaphase I, Metaphase I and Metaphase II, respectively. Tubulin and DAPI was used to label the spindle and DNA (green and blue, respectively). **(B)** Prophase-I arrested oocytes were co-microinjected with *Trim21* cRNA and either CCDC66 antibody or IgG. Oocytes were fixed at metaphase I and stained to detect CCDC66 (gray). Tubulin (green) and DAPI (blue) were used to label the spindle and DNA. **(C)** Relative CCDC66 intensity from B, two-tailed unpaired Students t-Test (***p < 0.001). **(D)** Quantification of percentage of polar body extrusion (PBE). **(E)** Percentage of aneuploid Metaphase II eggs after knockdown of CCDC66 in young and old mouse oocytes (One-way ANOVA, * p <0.05; ** p < 0.01, *** p < 0.001). Number of oocytes examined: young IgG: 34, young CCDC66: 37, old IgG: 29, old CCDC66: 31. These experiments were repeated 3 times. Scale bars: 15 and 4 μm (insets).

To evaluate a requirement for CCDC66 in oocyte meiotic maturation, we depleted the protein using the Trim away strategy [33] and confirmed ∼95% depletion by subsequent immunocytochemistry (Fig. 5B, C). To determine the effect of CCDC66 depletion on meiotic progression and meiosis I chromosome segregation, we calculated the percentage of oocytes that extruded polar bodies (PBE) and percentage of aneuploid Metaphase II eggs, respectively. In reproductively young mice (6-10 weeks of age, equivalent to ∼20 y of human age [42]), 73.18% of control-injected oocytes extruded a polar body. This rate decreased significantly to 66.16% in the CCDC66 depletion group (p < 0.05) (Fig. 5D). In oocytes from young mice, the average rate of aneuploidy in Metaphase II eggs was 2.56% in the control group and increased significantly to 13.24% in the depletion group (p < 0.05). Therefore, decreased expression of CCDC66 increases the chances of chromosome segregation errors during meiosis I in oocytes from reproductively young mice.

Elevated egg aneuploidy is associated with advanced maternal age (> 35 y) but some women experience higher egg aneuploidy rate at younger than average ages. To evaluate the interplay between genetics and maternal age, we also conducted the PBE and aneuploidy rate assessment experiments in reproductively older mice (9 m, equivalent to ∼38 y in humans [42]). Control-injected oocytes from older mice had a reduced PBE rate (66.59%) compared to oocytes in the young control-injected group (73.18%). Furthermore, depletion of CCDC66 also significantly reduced PBE compared to older oocyte controls (57.07% vs 66.59%, respectively; p < 0.05) (Fig. 5D). Similar to having an age-related reduction in PBE rate, control-injected oocytes from reproductively old mice had an elevated aneuploidy incidence (8.83%). Depletion of CCDC66 in oocytes from old mice had a more severe phenotype with a higher incidence of aneuploidy compared to controls (24.85%, p < 0.01). Furthermore, oocytes from 9-month-old mice were statistically more likely to be aneuploid when CCDC66 was depleted than oocytes from young mice. Taken together, these data demonstrate that decreased expression levels of CCDC66 is associated with increased egg aneuploidy rates, a phenotype which becomes more severe with reproductive aging.

## Discussion

The key to reproductive success lies in faithful chromosome segregation in meiosis to create a euploid zygote upon fertilization [1, 43]. The error-prone nature of meiosis often results in low quality gametes, leading to spontaneous abortion or infertility [3, 4, 44]. Recent studies suggest oocyte meiotic maturation is susceptible to dysregulation by maternal genetic variants that contribute to infertility, such as *CEP120* and *AURKB* (reviewed in [43, 45, 46]). These maternal genetic variants are strong candidates for clinical validation as predictive biomarkers of IVF outcomes. Identifying and validating additional genetic variants will contribute to a complete panel of infertility biomarkers. This can be used to complement existing clinical approaches to infertility, and genetic evaluations as the prognostic indicator of conception success could substantially improve pregnancy outcomes.

A major hurdle in identifying aneuploidy biomarkers is the lack of patient samples with both egg aneuploidy phenotypes and genome sequencing information. To overcome this limitation, we developed an integrated method for analyzing PGT-A data and illustrated the utility of these data for maternally-derived aneuploidy studies. We show that by leveraging the power of imputation and GLs, even ulc-WGS data are sufficient to identify common variant associations with aneuploidy risk, especially when aggregating sibling embryo sequences from the same patient. We discovered one novel locus associated with aneuploidy on chromosomes 3. Further eQTL analysis suggests that *CCDC66* is a novel candidate gene for embryo aneuploidy risk.

Through functional studies, we found that CCDC66 is important for the completion of meiotic progression and the production of euploid eggs. In mouse oocytes, the protein is expressed at all meiotic stages, and we observed a significant reduction of PBE in young and old mice after depleting endogenous CCDC66. Depletion of the protein also increased the incidence of aneuploidy, a phenotype that is exaggerated in aged mice. When the age and aneuploidy rate interaction was included as a co-variate in our association analysis, it did not show significant association with the aneuploidy rate variation. However, our limited sample size might have contributed to the result. In mitotic cells, CCDC66 function indicates that it is a microtubule-associated protein that localizes to centrosomes, centriolar satellites, and the primary cilium throughout the cell cycle [40, 41]. To our knowledge, this study is the first research on the function of CCDC66 in meiosis. Additional studies are needed to better understand its function in both mitosis and meiosis.

Our current study has a few limitations. First, in addition to errors of maternal meiotic origin, aneuploidy detected by PGTA could also arise from chromosome mis-segregation during early embryonic mitotic divisions. These mitotic errors could cause mosaicism in the embryos and potentially confound the meiotic aneuploidy phenotype of interest [47–50]. To circumvent this limitation, we recently developed a haplotype-based approach to isolate the subset of aneuploidies with characteristic signatures of meiotic error [20]. In the future when the sample size is sufficiently large, we can apply this method to disentangle the genetic underpinnings of mitotic versus meiotic errors. Analysis of these sub-phenotypes will allow us to evaluate whether certain alleles predispose to meiotic errors, mitotic errors, or both. Second, to increase the sequencing coverage, we combined embryo biopsy sequences from the same IVF cycle. Genetically, these embryos are equivalent of full siblings and the combined sequences contain genomic variation from both maternal and paternal genomes. Therefore, some parts of the genome could be tetraploid, rather than diploid. However, given the low coverage in the combined samples (median coverage 0.056×), we expect most of the sites are not affected. Third, using the PGT-A data from embryos, our analysis does not preclude paternal risk factors. Because embryonic aneuploidy is primarily derived from mistakes in female meiosis [51], we expect most of the association signals will be maternal risk factors. Nevertheless, functional studies in model organisms, where we knock-in mutations to mimic the human genetic condition, can help elucidate the role of the candidate genes in relation to their parental origin.

## Conclusion

Sufficient large sample size is fundamentally important in addressing biological questions in population and medical genetics. Large low-coverage sequencing datasets have become more accessible for analyses, as costs of sequencing continue to plummet. Given the same sequencing depth, low-coverage sequencing of many individuals tends to be more powerful than deep sequencing of fewer individuals [52, 53]. Recent studies have demonstrated the application of low-coverage sequencing data in GWAS [18, 54], polygenic risk score calculation [55], and population genomics [56, 57]. In addition, computational tools that are specialized for low-coverage sequencing data are also being actively developed [27, 58, 59]. These developments allow for future applications of low-coverage sequencing data.

Recently, a large number of ulc-WGS data have been generated from different sources, such as PGT-A [60], non-invasive prenatal testing (NIPT) [18], cell-free DNA (cfDNA) [61], and off-target sequencing reads from targeted sequencing experiments [62]. These sequences have not been fully investigated due to the difficulties in interpreting the sparse genotype observations. Our results show that when applied to large datasets, global patterns emerge even at the very low depth of coverage and can provide insight into the biological origins of aneuploidy. We believe that our method, with the consideration of genotype uncertainty in a probabilistic framework, will be applicable to other ulc-WGS datasets and will help improve the overall utility of the ulc-WGS data in the genetics field.

## Data Availability

All data produced in the present work are contained in the manuscript.

## List of abbreviations

IVF: *in vitro* fertilization
PGT-A: preimplantation genetic testing for aneuploidy
ulc-WGS: ultra-low-coverage whole-genome sequencing
GWAS: genome-wide association studies
PC: principal component
GL: genotype likelihood
MAF: minor allele frequency
GLM: generalized linear regression model
SNP: single nucleotide polymorphism
FDR: false discovery rate
MEM: minimum essential medium
CZB: Chatot, Ziomek, and Bavister
PFA: paraformaldehyde
PBS: phosphate-buffered saline
DAPI: 4, 6-Diamidino-2-Phenylindole, Dihydrochloride
eQTLs: expression quantitative trait loci
PBE: extruded polar bodies
NIPT: non-invasive prenatal testing
cfDNA: cell-free DNA

## Declarations

### Ethics approval and consent to participate

Not applicable

### Consent for publication

Not applicable

### Availability of data and materials

The summary statistics of the GWAS variants with p < 1e^−3^ is available in Table S1 and has also been submitted to the GWAS catalog (https://www.ebi.ac.uk/gwas/) (accession number pending). The analysis codes are available at https://github.com/JXing-Lab/PGTA_aneuploidy.

### Authors’ contributions

Conceptualization: R.C.M., J.X.; Investigation: S.S., M.A., D.A., M.E.H., C.R.W., M.D., A.B., M.V., M.K-J., R.C.M., K.S., J.X.; Data curation: D.A., M.E.H., C.R.W., M.V., M.K-J.; Formal analysis: S.S., M.A., D.A., R.C.M., K.S., J.X.; Initial draft: S.S., M.A., J.X.; Supervision: R.C.M., K.S., J.X.; All authors reviewed the manuscript and agreed to the published version of the manuscript.

## Acknowledgements

We thank the GTEx project for providing the eQTL data in the GTEx Portal. The GTEx Project was supported by the Common Fund of the Office of the Director of the National Institutes of Health, and by NCI, NHGRI, NHLBI, NIDA, NIMH, and NINDS.

